# Genome-wide postnatal changes in immunity following fetal inflammatory response

**DOI:** 10.1101/19000109

**Authors:** Daniel Costa, Núria Bonet, Amanda Solé, José Manuel González de Aledo-Castillo, Eduard Sabidó, Ferran Casals, Carlota Rovira, Alfons Nadal, Jose Luis Marin, Teresa Cobo, Robert Castelo

## Abstract

The fetal inflammatory response (FIR) increases the risk of perinatal brain injury, particularly in extremely low gestational age newborns (ELGANs, < 28 weeks of gestation). One of the mechanisms contributing to such a risk is a postnatal intermittent or sustained systemic inflammation (ISSI) following FIR. The link between prenatal and postnatal systemic inflammation is supported by the presence of well-established inflammatory biomarkers in the umbilical cord and peripheral blood. However, the extent of molecular changes contributing to this association is unknown. Using RNA sequencing and mass spectrometry proteomics, we profiled the transcriptome and proteome of archived neonatal dried blood spot (DBS) specimens from 21 ELGANs. Comparing FIR-affected and unaffected ELGANs, we identified 782 gene and 27 protein expression changes of 50% magnitude or more, and an experiment-wide significance level below 5% false discovery rate. These expression changes confirm the robust postnatal activation of the innate immune system in FIR-affected ELGANs and reveal for the first time an impairment of their adaptive immunity. In turn, the altered pathways provide clues about the molecular mechanisms triggering ISSI after FIR, and the onset of perinatal brain injury.

## Introduction

Intraamniotic infection (IAI), one of the main causes of spontaneous preterm birth, can lead to a maternal (MIR) and a fetal (FIR) inflammatory response, which can be detected in placental tissues^1^ (Supp. Table S1). FIR can damage the fetus presumably by triggering a local (e.g., lungs, skin) and systemic inflammation, which can lead to further distant organ injury, such as perinatal brain damage^2,3^. Understanding the molecular dynamics of FIR and its downstream alterations before and after birth is thus crucial to developing diagnostic and therapeutic strategies that improve the clinical outcome of extreme premature infants.

In a previous work, we found that FIR triggers a broad and complex transcriptional response in umbilical cord (UC) tissue^4^, which others have shown to correlate with cognitive impairment at 10 years of age^5^. The molecular and cellular alterations induced by FIR persist after birth^6,7^ and interact with postnatal inflammation-initiating illnesses^8^. This interaction ultimately leads to what is known as an intermittent or sustained systemic inflammation (ISSI)^9,10^. ISSI is strongly associated with perinatal brain injury and developmental disorders in ELGANs^10^ (Extremely Low Gestational Age Newborn, i.e. birth before the 28 week of gestation). The joint presence of placental inflammation and ISSI appears to increase the risk of perinatal brain injury, highlighting the important contribution of postnatal systemic inflammation to that risk^11,12^. However, little is known about the extent of postnatal molecular changes associated with FIR, which can contribute to understanding the link between FIR and ISSI and consequently provide an array of candidate biomarkers and therapeutic targets for perinatal brain injury.

Here, we analyze the transcriptome and proteome of archived dried blood spot (DBS) specimens from ELGANs and identify RNA and protein expression changes in whole blood that are significantly associated with FIR. These molecular changes provide, with an unprecedented level of resolution, a snapshot of the pathways participating in ISSI as a result of the exposure to FIR.

## Results

### Clinical characteristics of ELGANs and molecular profiling of DBS samples

The samples analyzed in this study were from 21 ELGANs (8 females and 13 males), who met the eligibility criteria (see Methods). Histological acute chorioamnionitis was diagnosed in 15 cases. MIR was in stage 1 in two cases, stage 2 in one case, and stage 3 in 12 cases. Ten of the 12 cases with stage 3 MIR showed FIR: four in stage 1, five stage 2, and one stage 3 (Supp. Fig. S1). Other clinical characteristics and outcomes of ELGANs are presented in Table 1, separately for those with (n=10) and without FIR (n=11).

Following the standard protocols at the hospital, all analyzed infants received antibiotic treatment. All but three mothers of FIR-affected ELGANs received at least two doses of a complete course of antenatal steroids, which in our study is compatible with the null hypothesis of no association with FIR (Table 1). None of the infants of the cohort received postnatal treatment with glucocorticoids. FIR-affected ELGANs presented a higher frequency of microbial invasion of the amniotic cavity (89% vs 29%, two-tailed Fisher’s exact test *P*=0.035), higher levels of interleukin-6 (IL-6) in amniotic fluid (AF) (*t*-test *P*=0.034) and lower levels of AF glucose (*t*-test *P*=0.018), than unaffected ones. The maternal blood C-reactive protein (CRP) levels at admission were also significantly higher in FIR-affected ELGANs (*t*-test *P*=0.041). Our data are compatible with no differences in neonatal morbidities between FIR-affected and unaffected ELGANs, although a non-significant higher frequency of neonatal sepsis and intraventricular hemorrhage was observed in FIR cases. Mean gestational age (GA) and birth weight were similar between the two groups, averaging 26 weeks and 860 g, respectively. Such an extremely low birth weight, with a lowest value of 580 g and up to 8 neonates under 750 g, precludes a standard blood extraction after birth for the purpose of molecular profiling in research. For this reason, DBS samples constitute a minimally invasive option to profile the transcriptome and proteome in peripheral blood from ELGANs.

**Table 1.** Clinical characteristics of ELGANs

| Clinical characteristics | No FIR (n=11) | FIR (n=10) | P value |
| --- | --- | --- | --- |
| *Baseline maternal characteristics* |  |  |  |
| Nulliparity: n, (%) | 9 (81.8) | 5 (50.0) | 0.183 |
| Preterm labor: n, (%) | 3 (27.3) | 3 (30.0) | 1.000 |
| Preterm, pre-labor rupture of membranes: n, (%) | 1 (9.1) | 4 (40.0) | 0.149 |
| Cervical insufficiency: n, (%) | 2 (18.2) | 3 (30.0) | 0.635 |
| Placenta abruption: n, (%) | 4 (36.4) | 0 (0.0) | 0.090 |
| Preeclampsia: n, (%) | 1 (9.1) | 0 (0.0) | 1.000 |
| Complete course (at least 2 doses) of antenatal steroids: n, (%) | 11 (100) | 7 (70.0) | 0.090 |
| Maternal CRP concentration at admission: (mg/dL), mean±SD | 2.3 ± 3.4 | 5.8 ± 3.8 | 0.041 |

| *Biomarkers of intra-amniotic infection and inflammation* |  |  |  |
| --- | --- | --- | --- |
| MIAC: n, (%) | 2 (28.6)<br>4 missing obs. | 8 (88.9)<br>1 missing obs. | 0.035 |
| Concentration of IL-6 in AF: (pg/mL), (Q1, Q3) | 21,153.5<br>(6,695.50,<br>35,975.75)<br>5 missing obs. | 112,271.5<br>(34,968.07),<br>162,866.25)<br>2 missing obs. | 0.034 |
| Concentration of glucose: (mg/dL), (Q1, Q3) | 23.6 (10, 37)<br>4 missing obs. | 2.4 (0, 2)<br>1 missing obs. | 0.018 |
| Clinical chorioamnionitis: n, (%) | 0 (0.0) | 3 (30.0) | 0.090 |
| Histological chorioamnionitis: n, (%) | 5 (45.5) | 10 (100.0) | 0.012 |
| *Perinatal outcomes* |  |  |  |
| Cesarean section: n, (%) | 8 (72.7) | 3 (30.0) | 0.086 |
| Male: n, (%) | 7 (63.6) | 6 (60.0) | 1.000 |
| GA: (wk), mean±SD | 26.5 ± 1.3 | 26.2 ± 1.2 | 0.489 |
| Birth weight: (g), mean±SD | 841 ± 138 | 880 ± 159 | 0.552 |
| Respiratory distress syndrome: n, (%) | 5 (45.5) | 5 (50.0) | 1.000 |
| Patent ductus arteriosus: n, (%) | 6 (54.5) | 6 (60.0) | 1.000 |
| Intraventricular hemorrhage: n, (%) | 1 (9.1) | 5 (50.0) | 0.063 |
| White matter disease: n, (%) | 1 (9.1) | 2 (20.0) | 0.586 |
| Retinopathy of prematurity: n, (%) | 5 (45.5) | 3 (37.5)<br>2 missing obs. | 1.000 |
| Necrotizing enterocolitis: n, (%) | 1 (9.1) | 0 (0.0) | 1.000 |
| Sepsis: n, (%) | 1 (9.1) | 5 (50.0) | 0.063 |
| DBS heel sampling days > 3: n, (%) | 4 (36.4) | 5 (50.0) | 0.678 |
| WBC count at birth: (10<sup>9</sup>/L), mean±SD | 8.6 ± 3.5 | 23.0 ± 15.6 | 0.017 |
| Postnatal WBC count within first 8 postnatal days: (10<sup>9</sup>/L), mean±SD | 13.2 ± 5.2 | 44.1 ± 13.4 | < 0.001 |
| Maximum ANC within first 8 postnatal days: (10<sup>9</sup>/L), mean±SD | 6.2 ± 4.3 | 29.0 ± 10.8 | < 0.001 |
| ALC at birth: (10<sup>9</sup>/L), mean±SD | 4.3 ± 1.2 | 6.7 ± 3.1 | 0.039 |
| Lymphocyte percentage at birth: (%), mean±SD | 54.3 ± 14.5 | 35.4 ± 16.3 | 0.012 |
| Maximum ALC within first 8 postnatal days: (10<sup>9</sup>/L), mean±SD | 6.0 ± 3.2 | 10.8 ± 4.1 | 0.009 |
| Maximum lymphocyte percentage within first 8 postnatal days: (%), mean±SD | 46.9 ± 17.1 | 25.4 ± 10.4 | 0.003 |
| Maximum CRP within first 8 postnatal days: (md/dL), mean±SD | 1.0 ± 2.2 | 3.5 ± 5.3 | 0.187 |

Total RNA sequencing of the 21 archived DBSs produced more than 500 million paired-end reads, which we transformed into a table of 25,221 gene-level summarized count expression profiles. Nearly half of these genes (11,279) showed reliable levels of expression in at least 6 of the 21 samples and were those selected for further analysis (see Methods). Figure 1a shows the distribution of sequencing depth across different genomic elements for each sample, where only about 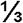 of the total sequencing depth derives from exonic reads. The small fraction of reads mapping to intergenic regions suggests no DNA contamination. DBS samples in this cohort were stored at room temperature for one to seven years until RNA extraction and sequencing. Over this period of time, degradation and fragmentation processes affect RNA integrity and we thus investigated whether, as observed in ancient DNA samples^14^, there is a detectable damage to the RNA due to cytosine deamination on such a short timescale (see Methods). Indeed, we found a significant linear relationship (r=0.8, *P* < 0.001) between the probability of a cytosine to thymine (C>T) substitution due to damage and the elapsed time between DBS extraction and RNA sequencing (Fig. 1b). Additionally, mass-spectrometry proteomics on DBSs produced 649 quantified protein expression profiles (Supplementary Data File 1), from which we selected 245 as being reliably expressed in at least 6 of the 21 samples (see Methods).

**Figure 1:**
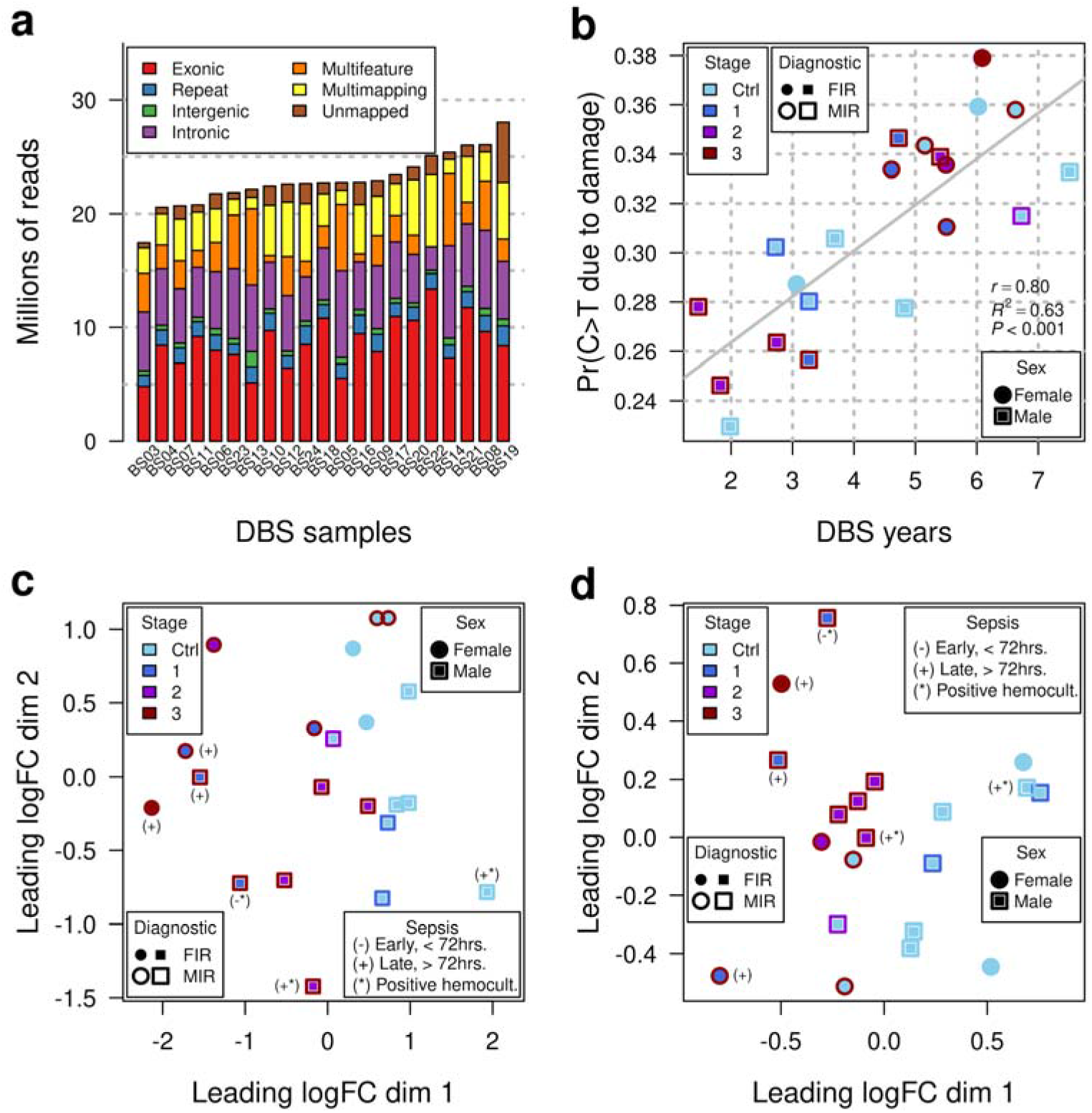
Transcriptomics and proteomics profiling of archived DBSs. (a) Sequencing depth per sample, broken by origin of sequence reads. (b) Probability of a C>T deamination due to damage, as function of DBS age in years. (c) Sample differences in terms of fold-changes between transcriptomic samples in base-2 logarithmic scale (logFC), projected along the x and y-axes. Distance between points is proportional to the dissimilarity between samples. (d) Same as (c) but calculated from proteomic samples.

Sample-level changes of transcriptomics data, projected in two dimensions (Fig. 1c), show that gene-level expression profiles derived from exonic reads capture the differences in placental inflammation throughout the different stages of MIR and FIR. Analogously, DBS protein expression profiles also show a clear separation between infants with and without MIR and FIR (Fig. 1d). In summary, transcriptomics and proteomics of archived DBS samples can produce a large catalog of molecular measurements that capture clinical features of the perinatal period.

### Neonatal DBSs contain gene and protein expression changes associated with FIR

The differential expression analysis of RNA-seq transcriptomic profiles between FIR and non-FIR exposed ELGANs identified 782 differentially expressed (DE) genes under an experiment-wide significance threshold of 5% false discovery rate (FDR) and a minimum 50%-fold change (Fig. 2a, Supp. Table S2). We replicated similar magnitude changes from DE genes on a logarithmic scale (r=0.75, R^2^=0.56, *P* < 0.001) in the analogous differential expression analysis of a DBS gene expression dataset^15^ from the US ELGAN cohort (Fig. 2b, Supp. Table S3 and Supp. Fig. S2 to S5), despite differences in sample preparation and gene expression profiling technology (see Methods). The hierarchical clustering of the 21 transcriptomic samples, using the 782 DE genes, leads to a perfect separation of the neonates who were affected by FIR, from those that were unaffected (Fig. 3).

**Figure 2:**
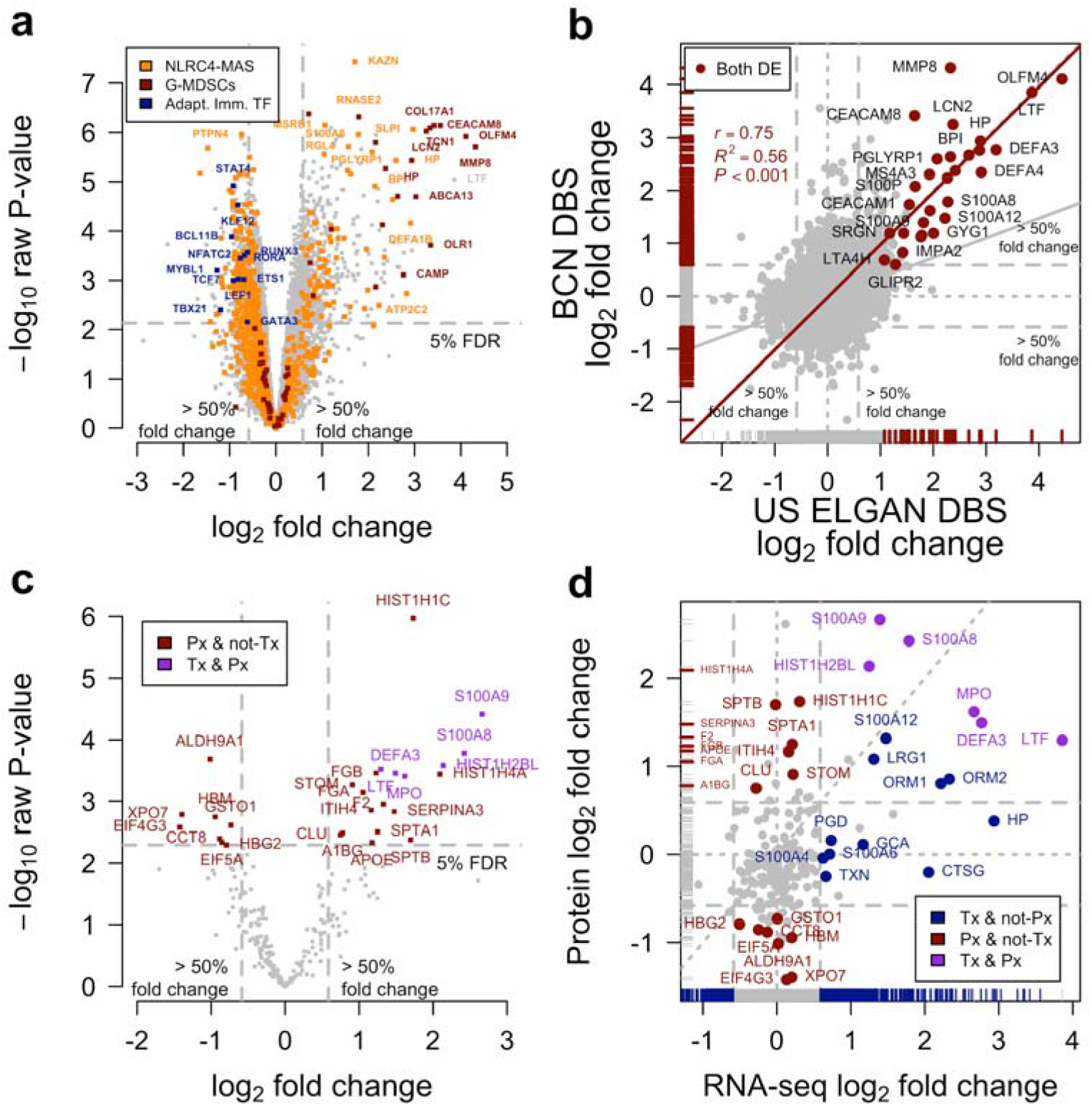
Differential expression analysis between FIR-affected and unaffected ELGANs, of transcriptomic and proteomic profiles from archived DBSs. (a) Volcano plot of transcriptomics data. Raw *P* values in negative logarithmic scale on the y-axis as a function of the log2-fold change on the x-axis. Orange dots highlight genes from a NLRC4-MAS signature^16^, red dots highlight genes from a G-MDSCs signature^17^ and blue dots highlight transcription factor coding genes involved in regulating adaptive immunity. (b) Fold changes of this study on the y-axis as a function of fold changes in data from the US ELGAN cohort in the x-axis. (c) Volcano plot of proteomics data. (d) Fold changes from proteomics data on the *y*-axis as function of fold changes from transcriptomics data on the *x*-axis.

**Figure 3:**
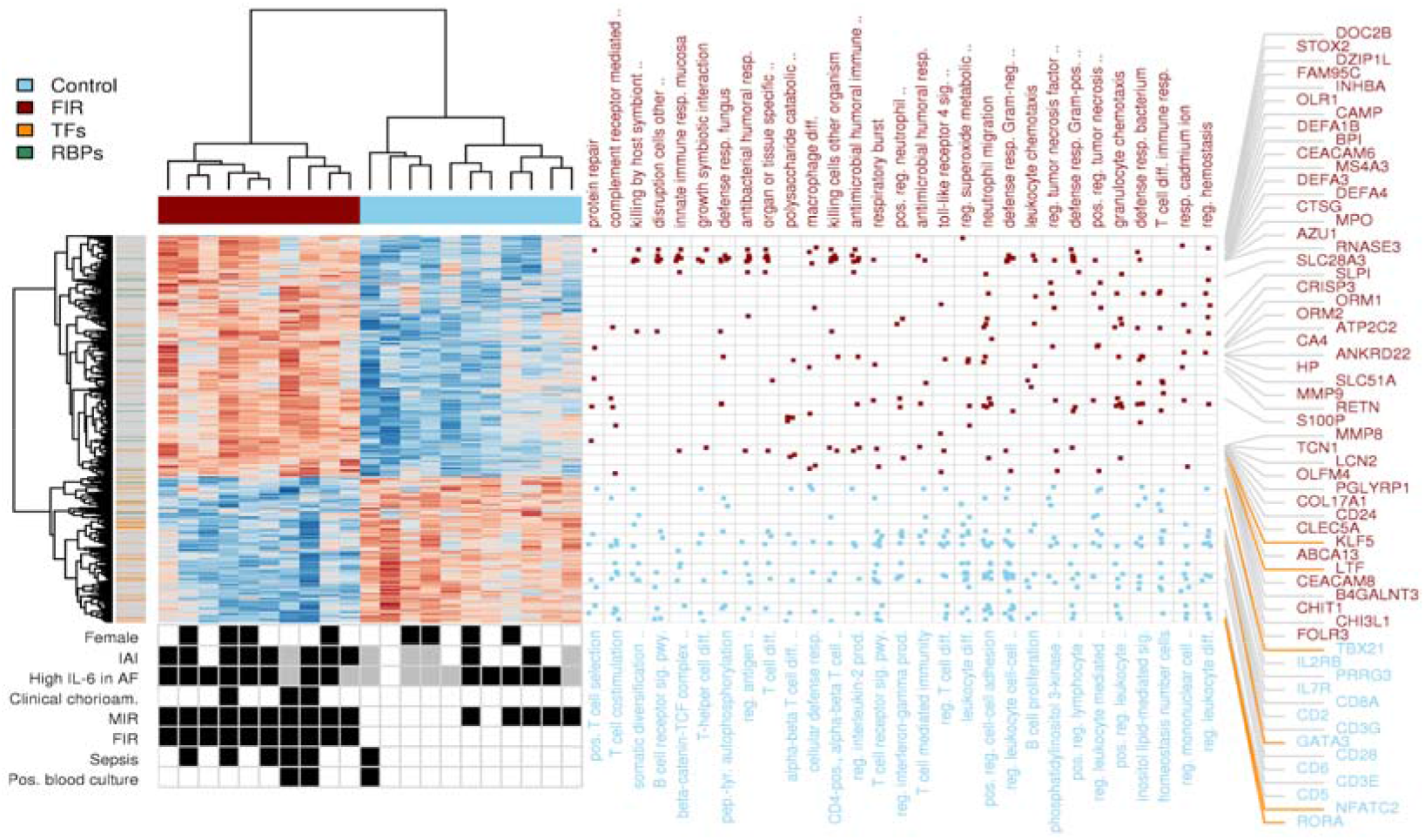
FIR gene expression signature and functional enrichment analysis. Heatmap of expression values for 782 DE genes with FDR < 5% and minimum 1.5-fold change between FIR-affected and unaffected ELGANs, obtained after removing FIR-unrelated variability. Dendrograms on the x and y-axes represent the hierarchical clustering of samples and genes, respectively. Leaves on the gene dendrogram are color coded according to whether genes encode for transcription factors (orange) or RNA-binding proteins (green), while those on the sample dendrogram are color coded according to FIR status, where red indicates samples derived from FIR-affected infants and blue unaffected. The right dot-matrix represents DE genes (y-axis) belonging to GO terms (x-axis) significantly enriched (FDR < 10% and odds-ratio -OR > 1.5) by upregulated (top) and downregulated (bottom) DE genes. The bottom-left dot-matrix represents the presence or absence of phenotypes, with grey indicating missing data.

A further question is whether there are RNA changes in peripheral blood across the different stages of MIR and FIR. Indeed, we found 58 genes with FDR < 5%, for which a combination of MIR and FIR stages, reflecting an increasing exposure to prenatal inflammation, led to a minimum increase of 50% in gene expression levels for each additional combined inflammatory stage (Supp. Fig. S6 and Table S4).

The differential expression analysis of proteomic profiles between FIR-exposed neonates and those who were unexposed identified 27 DE proteins at 5% FDR with a minimum 50%-fold change (Fig. 2c, Supp. Table S5). Figure 2d shows a comparison between log2-fold changes of the transcriptomics and proteomics analyses. Six of the 27 DE proteins are encoded by genes that are also DE by RNA-seq, while for 14 of them, their corresponding genes do not show fold-changes above 50% in the transcriptomics data. The remaining seven DE proteins do not have minimum levels of expression in the RNA-seq assay. Among them, F2, FGA, FGB and A1BG are secreted from the liver into the blood, and this may also be the case for SERPINA3 and APOE, which have a biased expression towards the liver. Overall, these results show that in peripheral blood from ELGANs there are sizable numbers of transcriptomic and proteomic changes associated with FIR that can be detected in archived DBS.

### FIR is associated with a postnatal activation of the innate immune system

Blood protein biomarkers have helped in the discovery of a postnatal systemic inflammatory hit following FIR^6^. However, the extent of such molecular changes was previously unknown. Our transcriptomic data from DBS samples show the postnatal upregulation in FIR of many genes involved in the innate immune response (Fig. 2a; Supp. Table S2). Such genes include those coding for pattern recognition receptors (PRRs, e.g., *NLRC4, CLEC4E, FPR1*), inflammatory transcription factors (e.g., *CEBPD, KLF5*), acute phase response proteins (*HP, ORM1*), as well as multiple inflammatory mediators including cytokines (e.g., *IL18, IL1R1, RETN*), calgranulins *(S100A8, S100A9,S100A12)*, chemokines (e.g., *CXCR1, CXCL1*), complement (e.g., *C5AR2, C3AR1)*, lipid mediators (e.g., *LTA4H, ALOX5AP, CYP4F3*), proteolytic enzymes (e.g., *MMP8, MMP9*) and adhesion molecules (e.g., *ICAM3, ITGAM*). Our proteomic data from DBS samples (Fig. 2c, Supp. Table S5) also shows protein overexpression in FIR of diverse inflammatory mediators of the innate immune system, such as the S100A8, S100A9, LTF, MPO and DEFA3 proteins. A functional enrichment analysis using the Gene Ontology (GO) database (Fig. 3) shows GO terms associated with the innate immune system, such as *innate immune response in mucosa* (OR=17.7) and *defense response to fungus* (OR=14.3), which were significantly enriched (FDR < 10%) in upregulated genes (Supp. Table S6; see Methods). We also observed a significant enrichment of DE genes in curated gene sets of the innate immune response, such as the InnateDB^18^ database (one-tailed Fisher’s exact P < 2.2 × 10^−16^, OR=3.2).

Moreover, we found that FIR upregulates the expression of the NLRC4 inflammasome and IL-1 family-related protein-coding genes (e.g., *NLRC4, PYCARD, IL18, IL1R1*), including a significant overlap (Fig. 2a, one-tailed Fisher’s exact P < 2.2×10^−16^, OR=10.4) of a transcriptional signature of the NLRC4-infantile-onset macrophage activation syndrome (MAS)^16^ with our set of 782 DE genes. Consistent with these results, we observed the upregulation of numerous genes involved in neutrophil biology and function (Supp. Table S7), significantly enriching (FDR < 10%, OR > 1.5) GO terms related to neutrophil activation^19^, such as *neutrophil migration* (OR=6.3) or *positive regulation of neutrophil chemotaxis* (OR=7.5); see Fig. 3 and Supplementary Table S6.

Previous studies of postnatal molecular changes associated with FIR in ELGANs^6,7,10^ were based on a handful of gene and protein biomarkers. The gene and protein expression changes described here constitute, to our knowledge, the most comprehensive description to date of the postnatal activation of the innate immune system in ELGANs affected by FIR. Our data suggest that FIR leads to a postnatal robust activation of the IL-18/IL-1 axis through the NLRC4-inflammasome that can induce neutrophil activation, contributing to a systemic inflammation.

### FIR is associated with the impairment of adaptive immunity and thymic function through the expansion of granulocytic myeloid-derived suppressor cells

The mechanism by which antenatal FIR leads to a postnatal systemic inflammation is unknown. Among several plausible alternatives, Dammann and Leviton (2014)^9^ suggested an alteration in the number of T cells and their regulation. A closer look at the hierarchical clustering of genes in Figure 3 reveals an enrichment of downregulated transcription-factor coding genes (one-tailed Fisher’s exact test *P* < 0.001, OR=2.9, Supp. Table S8) in FIR-affected ELGANs. Many of these genes are regulators of T cell development and activation, such as *TBX21, GATA3* and *RORA*, respectively involved in T-helper(h) fate of CD4+ T cells in Th1, Th2 and Th17 lineages (Fig. 2a). More generally, we found the downregulation of key genes associated with the IL-7 and IL-2 signaling pathways (e.g., *IL7R, BCL2, PIK3AP1, IL2RB, CD3E, CD3G*) involved in T cell development (Supp. Tables S2 and S9). The T cell receptor signaling pathway (e.g., *CD3E, CD3G, LCK, FYN, ZAP70, CD247*) and T cell differentiation co-stimulatory receptor genes (e.g., *CD28, ICOS, SLAMF1*) were also downregulated^20^. Likewise, the inhibitory and stimulatory genes *KLRC1, KLRC3, KLRC4* that belong to the natural killer (NK) cell lectin-like receptors of the NK gene complex, and genes involved in NK cell cytotoxicity, such as *PRF1, SH2D1B* genes, were also downregulated in FIR-affected ELGANs^20^. Interestingly, we observed a significant overlap of downregulated genes with an NLRC4-infantile-onset MAS signature (Fig. 2a, one-tailed Fisher’s exact P < 2.2×10^−16^, OR=21.5)^16^, which is associated with cytotoxic and regulatory T cell dysfunction.

Recent thymic emigrants consist of phenotypically and functionally immature T cells that have recently egressed from the thymus, which predominate in the fetus and newborns. These cells can be identified by their content of T cell receptor excision circles (TRECs)^21^, which are DNA fragments excised during the rearrangement of segments of T cell receptor genes in the thymus. TRECs are used as a measure of the number of T lymphocytes in newborns and, indirectly, as an indicator of a possible thymus dysfunction. We found downregulation of the genes coding for the PTK7 and S1PR1 proteins in FIR-affected ELGANs. PTK7 is a cell marker of CD4+ T cell recent thymic emigrants used for assessing thymic activity^22^, while the S1PR1 receptor expression in CD4+ T cells is required for their survival and thymic emigration^23^.

Consistent with these results, FIR-affected ELGANs had significantly lower levels of TRECs in the analyzed DBSs (P=0.0432; Fig. 4a, see Methods), a significantly higher postnatal neutrophil to lymphocyte ratio (P=0.002; Fig. 4b), and a significantly lower percent of all leukocytes that are lymphocytes at birth (P=0.012; Table 1) and during the first postnatal week^24^ (P=0.003; Fig. 4c).

**Figure 4:**
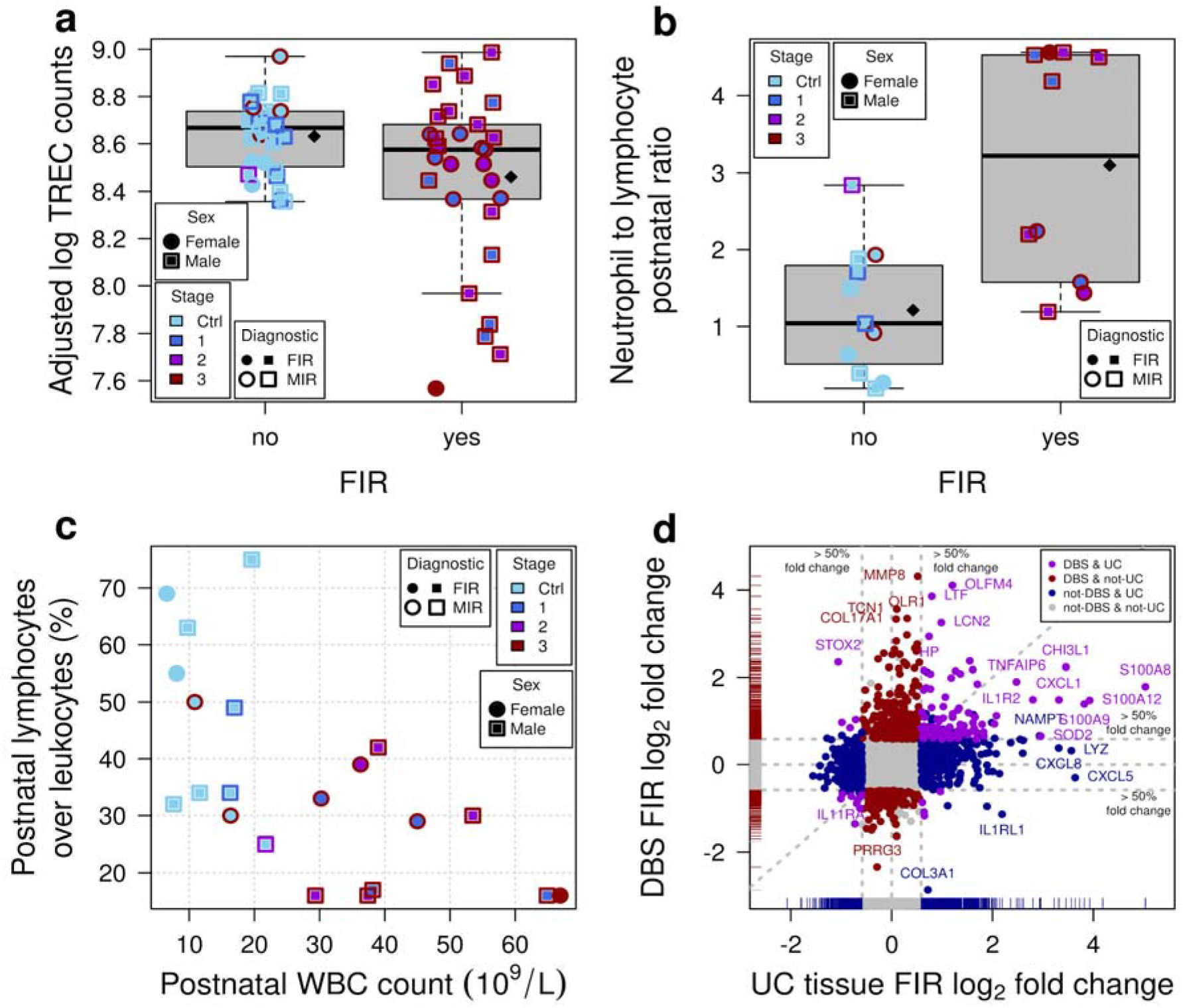
Association of FIR with clinical and molecular signatures. (a) TREC corrected fluorescent counts on logarithmic scale by FIR status, after adjusting for sex, maximum lymphocyte count during the first postnatal week and batch. Diamonds indicate mean values. There are multiple measurements per newborn. (b) Postnatal ratio of neutrophil absolute count to absolute lymphocyte count, during the first postnatal week, by FIR status. Diamonds indicate mean values. (c) Percent of all leukocytes that are lymphocytes during the first postnatal week on the *y*-axis as a function of the postnatal white blood cell count on the *x*-axis. (d) Fold changes in this study on the *y*-axis as a function of FIR fold changes in UC tissue on the *x*-axis.

Myeloid-derived suppressor cells (MDSCs) are immature myeloid cells that can develop under certain pathological conditions and are central regulators of immune responses. The hallmark of MDSCs is their immune suppressive activity, and according to their phenotype, MDSCs can be classified into monocytic(M)-MDSCs or granulocytic(G)-MDSCs. Postnatally, there is a high level of circulating MDSCs, predominantly G-MDSCs, which correlates with inflammatory markers of perinatal infection^25,26^. Neonatal MDSCs display reduced apoptosis and immunosuppressive activity after bacterial infection^27^. Indeed, these cells are potent inhibitors of adaptive immune responses, particularly of T and NK cells, by different mechanisms, including the production of reactive oxygen species (ROS)^28–30^, prostaglandin E2 (PGE2), and the depletion of L-arginine by the arginase 1 enzyme (ARG1)^31,32^. In our data, we found that the *ARG1* gene was 2.5-fold upregulated in FIR-affected ELGANs, as well as the *OLR1* gene, encoding the lectin-type oxidized LDL receptor 1 (LOX1), a marker of G-MDSCs^17^, which was over 10-fold upregulated. We also found a significant overlap (one-tailed Fisher’s exact P < 2.2e-16, OR=26.4) between FIR upregulated genes and a G-MDSC expression signature^17^ (Fig. 2a).

One of the mechanisms by which G-MDSCs suppress T cell responses is the release of ROS molecules. We found an upregulation of genes coding for proteins essential to ROS production such as *CYBB, NCF1, NCF4, MPO, RAB27A*, as well as genes coding for antioxidant enzymes (e.g., *SOD2, TXN* and the Msr family *-MsrA,MsrB1, MsrB2, MsrB3)*. The Msr gene family encodes proteins of the methionine sulfoxide reductase system that reduces protein methionine oxidation, a post-translational alteration that correlates with oxidative stress^33^. In summary, our data suggest the expansion of G-MDSCs as a mechanism contributing to the impairment of adaptive immunity in ELGANs affected by FIR, and consequently, to a postnatal systemic inflammation sustained over time after birth.

## Discussion

We have described the largest catalog to date of postnatal blood transcriptomic and proteomic changes associated with FIR in archived DBSs from ELGANs. This catalog demonstrates that archived DBSs, collected for the newborn screening program, constitute a valuable source of genome-wide molecular information about the perinatal period. More importantly, we have shown that molecular changes in this catalog provide clues to the underlying relationships between FIR and ISSI, with common and specific alterations to each of these two conditions (Fig. 4d and Supp. Table S10).

Our data provide molecular evidence of the postnatal activation of NLRC4-inflammasome dependent mechanisms, which may contribute to the development of ISSI in FIR-affected ELGANs. After cytosolic detection of microbial and endogenous danger signals by NOD-like receptors, the NLRC4 inflammasome induces, via activation of caspase-1 (CASP1), the maturation and release of IL-1β and IL-18 cytokines, the bacterial clearance and the pyroptosis of immune cells. The release of interleukin-1 family cytokines such as IL-1β and IL-18 has potent effects on neutrophil recruitment, activation and lifespan^34^. However, the NLRC4-inflammasome hyperactivity can impair inflammation resolution and cause diverse auto-inflammatory diseases, such as the infantile-onset MAS syndrome and enterocolitis. These disorders are characterized by a huge persistent elevation of IL-18 in the blood^16^ as in neonatal sepsis^35^ and, in fact, we observed a 2-fold overexpression of the *IL18* gene in FIR-affected infants. Liang et. al (2017)^36^ reported recently the case of a 28-week preterm neonate with a *de novo* gain-of-function mutation in the *NLRC4* gene, who died of a fatal syndrome of excessive immune activation likely to have begun in utero. NLRC4 dependent mechanisms can also contribute to brain injury induced by cerebral ischemia, as has recently been described in a rodent-based model of stroke^37^. Interestingly, caffeine therapy for the apnea of prematurity, which reduces the incidence of moderate to severe neurodevelopmental disabilities in very low birth weight infants^38^, can also reduce NLRC4-inflammasome activation^39^.

The resolution of inflammation requires neutrophil arrest, death and removal^40^. However, a high and persistent release of the proinflammatory cytokines IL-18 and IL-1β after NLRC4 inflammasome hyperactivation may induce the continued expansion, migration and activation of neutrophils. Moreover, molecular alterations in neonatal neutrophils, such as a reduced expression of Fas receptors, can lead to delayed neutrophil apoptosis^41,42^ and the persistence of proinflammatory responses^43^. In FIR-affected ELGANs, we found a significantly higher neutrophil to lymphocyte ratio (Fig. 4b), the overexpression of genes involved in neutrophil biology, such as *S100A8, S100A9, IL18, MMP8* and *OLFM4*, and the underexpression of gasdermin B genes (Supp. Table S2). Calgranulins S100A9 and S100A8, and IL-18 can inhibit neutrophil apoptosis^44,45^ and neutrophils can evade pyroptosis, which is mediated by gasdermin family proteins^46^. Furthermore, the robust production of calgranulins and the activation of the IL-1/IL-18 axis can trigger a neutrophil-associated inflammation-boosting loop binding to PRRs and triggering downstream inflammatory signaling pathways, just as in auto-inflammatory disorders^47^. Among children with septic shock, elevated *MMP8* and *OLFM4* blood gene expression levels, which are markers of neutrophil activation, are associated with higher rates of mortality and organ failure^48,49^. Similarly, other studies have also shown that preterm neonates with abnormal neuroimaging studies had increased blood neutrophil activation markers^50,51^.

Chorioamnionitis can lead to an inflammation-induced immunosuppression similar to the sepsis-induced immunosuppression^52,53^. This chorioamnionitis-induced immunosuppression includes the depletion of B and T lymphocytes in spleen^54^, thymic involution^55^, endotoxin hyporesponsiveness^56^ and lower CXCL8-producing CD4 and CD8 T cells^57^. In umbilical cord blood monocytes from very preterm newborns (GA 29-32 wk.) affected by chorioamnionitis, it has been described the downregulation of genes involved in adaptive immunity upon monocyte stimulation with Staphylococcus epidermidis^58^, the most common cause of neonatal late-onset sepsis. The exposure of umbilical cord blood monocytes to chorioamnionitis has been also shown to alter the histone modification landscape and reduce the immune response to a second inflammatory stimulus with lipopolysaccharides^59^. The molecular events underlying this chorioamnionitis-induced immunosuppression are, however, not well characterized in extremely preterm newborns, particularly after birth. In our data, the analysis of downregulated genes provides for the first time molecular evidence of the postnatal inhibition of adaptive immune responses, particularly T cell, in FIR-affected ELGANs (Fig. 2a and 3). This evidence is consistent with the observed relationships between TRECs and FIR (Fig. 4a), as well as the lymphocyte percent among ELGANs with and without FIR and MIR (Fig. 4c). The reduction of TRECs and the downregulation of the *PTK7* and *S1PR1* genes suggest a thymic involution associated with FIR^60^, which may lead to a relative increase in proinflammatory CD31-CD4+ T cells producing TNF^61^. Perhaps that is why thymic involution has also been associated with increased risk of brain damage in ELGANs^62^.

Previous findings are also consistent with our data because we found a significant overlap between FIR DE genes and a published G-MDSC signature^17^(Fig. 2a), as well as the underexpression of the *LCK* and *ZAP70* genes and the overexpression of *S100A8, S100A9* and *LTF* genes and protein products in FIR-affected ELGANs (Fig. 2d). In newborns, the LTF protein triggers a G-MDSCs T cell suppressive phenotype, mediated by the secretion of S100A8, S100A9 and PGE2^63^, which repress the expression of LCK and ZAP70 in CD4+ T cells, impairing their activation^29,30,64^. Overall, our results suggest that after an initial prenatal pro-inflammatory-driven response evident in UC tissue (e.g., funisitis) and UC blood (IL-6, IL-1β, CXCL8), FIR is followed by an intense neonatal innate immune activation mediated by the NLRC4-inflammasome hyperactivity. As a result of these events, FIR-affected ELGANs display a robust neutrophil and G-MDSC expansion during the first postnatal days, which can lead to adaptive immune suppression, thymic involution, delayed neutrophil apoptosis and the onset of an auto-inflammatory loop, contributing to ISSI and consequently increasing the risk of perinatal brain injury.

An important feature of G-MDSCs is the production of ROS molecules, which can further lead to oxidative stress and damage. Preterm newborns have a reduced antioxidant capacity that increases the risk of ROS-induced damage, which is an important factor in numerous diseases in premature infants, such as perinatal brain damage^65^. In fact, ELGANs are born with deficient levels of selenium^66^, a cofactor for the MsrB and thioredoxin enzyme activity required for proper antioxidant function. We found overexpression of genes coding for antioxidant enzymes (e.g., Msr family genes) and a 10-fold overexpression in FIR of both the *OLR1* gene, which encodes LOX1, a marker of G-MDSCs^17^, and the gene coding for the heat shock protein HSPA1A. HSPA1A can activate LOX1 in UC neutrophils in response to in vitro exposure to microbial peptidoglycan^67^, leading to the downstream production of ROS. The LTF protein, which we found to be significantly overexpressed in both transcriptomics and proteomics data, can protect newborns from oxidative stress and damage by attenuating inflammatory responses and controlling oxidative cell injury induced by innate immune activation^68^.

Using well-established biomarkers, it has been shown that antenatal and postnatal inflammation, the so-called two-inflammatory hit, increases the risk of perinatal brain damage in preterm newborns^11^. Here, we found the overexpression of multiple genes encoding proteins that can be associated with the onset of perinatal brain damage in FIR-exposed newborns, such as LCN2, CXCL1, CXCR2, S100A8 and S100A9, as well as histone-related proteins (e.g., HIST1H2BL). LCN2 is an iron-binding protein contained in neutrophil granules that is associated with neuroinflammation^69^. Using an animal model of chorioamnionitis, Yellowhair et al. (2018)^70^ found that chorioamnionitis-induced fetal brain damage was associated with increased expression of CXCL1 and CXCR2 in placental and fetal brain, and with an elevated number of cerebral CXCR2+ neutrophils. With respect to calgranulins, it has been observed that the brain expression of S100A8 and S100A9 proteins was increased in the brain of patients dying of sepsis; specifically, S100A9 expression was required for the cerebral recruitment of neutrophils and microglia activation in an animal model of sepsis^71^.

Extracellular histones can also act as damage-associated molecular patterns, such as calgranulins, activating inflammatory signaling pathways and inducing multiple organ-specific damage^72^. In an animal study, histone H1 was found to induce proinflammatory responses in microglia and astrocytes^73,74^. We also found that both the gene and protein expression of S100A9, S100A8 and HIST1H2BL, were significantly higher in FIR-affected ELGANs, while our proteomics data also revealed the significant overexpression of HIST1H4A and HIST1H1C (Fig. 2c,d). In summary, our findings provide new insights into the molecular mechanisms that trigger ISSI after FIR and the onset of perinatal brain injury in ELGANs.

## Methods

### Study design

We performed a retrospective chart review of extreme preterm births (< 28 weeks of gestation) at the *Hospital Clínic de Barcelona* admitted to the neonatal intensive care unit (NICU) between January 2010 and October 2016. Two hundred thirty-four cases born alive were found. From this initial cohort, we excluded newborns from multiple pregnancies, outborn deliveries and those who presented major congenital anomalies, received a blood transfusion before collecting the DBS, had no histopathological examination of the placenta and umbilical cord, or whose DBS was collected later than 8 days after birth, most likely presenting molecular alterations unrelated to intrauterine conditions. To further avoid molecular alterations caused by severe organ dysfunction unrelated to FIR, we also excluded newborns who either died in the delivery room or within the first week after birth. We attempted to maximize the number of individuals with available biomarkers of intra-amniotic infection and inflammation in placenta and AF, while recruiting a similar number of FIR and non-FIR cases within a total maximum of 25, which as our results show, is sufficiently large to obtain a sizeable number of transcriptomic and proteomic changes. While this sample size may apparently be insufficient to enable a proper adjustment by relevant covariates, the high-dimension of the molecular data allows one to circumvent that limitation by using surrogate variable analysis^75^, as described later in this section. A final number of n=21 infants met the eligibility criteria and their families provided written consent to use their DBS samples for this study. At the time of processing these DBS samples, they had been stored at room temperature by the Catalan newborn screening program for between 1 and 7 years. The study protocol was approved by the Institutional Review Board from the *Hospital Clínic de Barcelona* (September 16^th^, 2016; ref. HCB/2016/0713).

### Clinical outcomes

GA at delivery was calculated according to the first-trimester ultrasound examination. Preterm birth was classified by the clinical presentation: threatened preterm labor, cervical insufficiency, preterm pre-labor rupture of membranes, placenta abruption, and preeclampsia. The laboratory results of AF cultures (genital mycoplasma, aerobic and anaerobic) and IL-6 levels, as well as the mother’s blood CRP concentration, white blood cell (WBC) count and absolute neutrophil count (ANC) at admission were also recorded; see Table 1. Clinical chorioamnionitis was defined following the criteria of Gibbs, which includes presence of fever and any two of the following: maternal leukocytosis, maternal tachycardia, fetal tachycardia, uterine tenderness and foul-smelling vaginal discharge^76^. Slides from the placenta and umbilical cord were independently reviewed by two experienced pathologists blinded to experimental results and previous diagnosis. Maternal (MIR) and fetal (FIR) reactions were staged and graded according to published criteria^1,77^. Cases read discrepantly by the two pathologists were simultaneously re-reviewed on a two-headed microscope in order to reach a consensus (Supp. Table S1 and Supp. Fig. S1). The annotated neonatal morbidity included: respiratory distress syndrome (RDS); intraventricular hemorrhage (IVH); white matter disease (WMD); retinopathy of prematurity (ROP); sepsis; necrotizing enterocolitis (NEC); patent ductus arteriosus (PDA); and laboratory investigations at birth, and during the first 8 days after birth (CRP concentration, WBC and ANC). Hypothesis tests on clinical variables used in Table 1 include two-tailed Fisher’s exact test for categorical variables and *t*-tests for those that were continuous.

### RNA extraction, library preparation and sequencing

We used a scalpel to cut a surface of approximately 25 square millimeters from DBS samples in Guthrie cards, using a different scalpel blade for each sample to avoid contamination. RNA was then extracted using the illustra™ RNAspin Mini Isolation Kit (GE Healthcare) following the supplier’s protocol and eluting the sample in a final volume of 30 μl. The quality and quantity of the RNA was assessed with an RNA Pico chip in an Agilent 2100 Bioanalyzer equipment. Next, globin mRNA and ribosomal RNA were removed using the Globin Zero Gold rRNA Removal Kit (Illumina), and its performance was checked with an RNA Pico chip in an Agilent 2100 Bioanalyzer equipment. Libraries were prepared using the NebNext UltraTM II Directional RNA Library Prep Kit (New England Biolabs), following the specific protocol for rRNA depleted FFPE RNA and using 12 PCR cycles for library amplification. Libraries were validated and their concentration was measured with a High Sensitivity DNA chip in an Agilent 2100 Bioanalyzer. Finally, we prepared a pooled library with a normalized concentration for each sample. The final concentration of the pool was measured by Real-Time PCR using the NGS Library Quantification Kit (Takara). We loaded a final concentration of 1.9 pM into a NextSeq-500 High Output run with 2×75 cycles using 4 sequencing lanes per library. The resulting 21×4=84 paired-end FASTQ files have been deposited to the European Genome-Phenome Archive with the study identifier EGAS00001003635.

### Pre-processing and differential expression analysis of transcriptomics RNA-seq data

We performed quality control on the 84 raw paired-end reads using FastQC and did not detect artifacts in the sequence data. We did observe, however, that one of the samples, BS13, was sequenced at about 4 times more depth than the rest of the samples. The reason for this higher depth was a wrong normalization step during library preparation that led to a higher concentration of RNA in that sequenced library. To adjust for this bias in sequencing depth we downsampled uniformly at random the paired-end reads from the library to 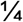 of its original depth.

Raw paired-end reads in FASTQ files, including the downsampled version of BS13, were aligned to the GRCh38 version of the reference human genome, without alternate locus scaffolds (GCA_000001405.15) and including human decoy sequences from hs38d1 (GCA_000786075.2), using STAR^78^ version 2.6.0c with default parameters except for **–** `peOverlapNbasesMin 10` and `--sjdbOverhang 74`, producing an initial set of 84 BAM files. Aligned reads in BAM files were reduced to a table of counts of 25,221 genes by 84 samples using gene annotations from GENCODE v24 and the R/Bioconductor package *GenomeAlignments^79^* version 1.16.0, and its function summarizeOverlaps. We used specific arguments in the call to this function to restrict the count of genic reads to only those that fell entirely within the exonic regions and aligned to a unique site on the genome, to reflect library preparation protocols and to avoid counting reads without a matching pair or overlapping multiple features.

The rest of the analysis of transcriptomics data was based on the *edgeR^80^* and *limma^81^* pipelines, and a few other R packages from the Bioconductor project^82^ release version 3.7. A multidimensional scaling (MDS) plot of the samples showed a nearly perfect agreement between libraries from the same sample sequenced in each of the 4 different lanes. For this reason, we merged BAM files with aligned reads from the same sample and built a new table of counts of 25,221 genes by 21 samples, using the previously employed reduction procedure.

Considering that 6 out of the 21 infants were not diagnosed with MIR or FIR (Supp. Fig. S1), and following previously established recommendations^83^, we filtered out lowly-expressed genes by discarding those that did not show a minimum reliable level of expression of 10 counts per million reads of the smallest library size, in at least 6 samples. This filtering step led to a final table of counts of 11,279 genes by 21 samples.

We used TMM normalization^84^ to produce the MDS plot shown in Figure 1c, as well as to obtain gene-level robust dispersion estimates^85^ that helped to select the sex of the infant and surrogate variables calculated with SVA^75^, as covariates in the linear models employed for differential expression analysis. Finally, we used the limma-voom^86^ pipeline with quality weights, combined with quantile normalization, to conduct the differential expression analysis using linear models. The design matrix included the FIR status of the infants as a main explanatory variable and sex and surrogate variables, estimated with SVA, as covariates.

The differential expression analysis across different combined stages of inflammation in MIR and FIR (Supp. Fig. S6, Supp. Table S4) was based on our replacing the FIR status variable in the linear model, with numerical values for MIR and FIR. Specifically, we assigned a 0 when MIR and FIR were not present, 1 when MIR was present but FIR was not, 2 when MIR was stage 3 and FIR was stage 1 and, finally, 3 when MIR was stage 3 and FIR was stage 2 or 3.

### Pre-processing and differential expression analysis of transcriptomics microarray data

We obtained from Nigel Paneth and collaborators at the Michigan State University raw microarray gene expression data files from paired frozen and unfrozen DBS samples from nine neonates of the US ELGAN cohort, where two were affected by FIR and seven were not. The n=18 corresponding samples had been hybridized on a Agilent SurePrint G3 Human Gene Expression 8×60K Microarray Kit, and their analysis comparing the number of expressed genes between frozen and unfrozen samples was published by Wei *et al*. (2014)^15^. Batch processing information was derived from the scanning timestamp stored in raw data files as previously described^87^, creating a batch indicator variable that divided the samples into two different batches. The cross-classification of infants by FIR status and batch indicator showed no correlation between the primary outcome of our analysis, FIR status, and sample batch processing (Supp. Fig. S2 and S4). The MDS plot of the data shows the effect of storage temperature and batch processing, but not sex (Supp. Fig. S4).

We corrected background expression intensities using the “normexp” method in the function backgroundCorrect from the *limma* package^88^, and used a quantile normalization approach to obtain comparable gene expression distributions across the samples (Supp Fig. S3). We removed probesets without annotation to Entrez gene identifiers, and probesets annotated to a common Entrez gene, keeping the one with highest variability measured by interquartile range (IQR). We then discarded the Agilent positive control probes and retained initially the negative control probes to later estimate the fraction of expressed genes per sample. This resulted in an expression data set of 21,952 probesets in one-to-one correspondence with Entrez genes after discarding negative control probes, by n = 18 samples. We calculated the 95% quantile of the expression level distribution observed in negative control probes for each sample. As a function of the percentage of increase over the 95% quantile of negative probe expression (from 100% to 200% increasing by 10%), we found the number of expressed genes per sample separately by storage condition, which we later used for non-specific filtering based on minimum expression.

Using the software package *sva*^75^ and the top 1% most variable (IQR) genes, we estimated one surrogate variable covariate that captures variability unrelated to FIR, storage temperature and batch processing. For each gene, we defined a linear mixed-effects model where the expression profile of every gene is a linear function of FIR status (yes/no, main explanatory variable), storage temperature (frozen/unfrozen), batch processing, the estimated surrogate variable covariate and the infant as a random effect. We estimated array quality weights to down-weight samples of worse quality in a graduated way^89^ and to avoid discarding them. Using limma, we fitted this linear mixed-effects model, including the array quality weights, to each gene expression profile. We then calculated moderated t-statistics, squeezing the genewise residual variances towards a global trend, for the coefficient estimating the effect between FIR and non–FIR-affected infants, and their corresponding *P* values for the null hypothesis of no-differential expression (Supp. Fig. S5). We selected initially different subsets of DE genes at 5% FDR according to different thresholds of the non-specific filtering on minimum expression and, within each non-specific filter, FDR-adjusted *P* values were re-calculated. We found that a 110% cutoff yielded the highest number of DE genes and using this non-specific filter we readjusted the raw *P* values and obtained 33 genes that changed significantly at 5% FDR by 50% or more, between newborns affected and unaffected by FIR (Fig. 2b, Supp. Fig. 7b and Table S3).

### Mass spectrometry proteomics

#### Sample preparation for mass spectrometry

DBS samples were excised and collected in a 2 mL eppendorf, and extracted as described previously^90^. Briefly, each sample was soaked with 970 μl of a solution consisting of 758 μl of 25 mM ammonium bicarbonate, 113 μl of 10% (w/v) sodium deoxycholate and 99 μl of 5 mM tris(2-carboxytethyl)phosphine, which was incubated at 60 °C during 1h in constant agitation. Samples were alkylated with 10 μM iodacetamide (37°C, 30 min) and quenched with 10 μM dithiothreitol (37°C, 30 min). Samples were digested with trypsin (40 ng/μl) overnight at 37 °C. Digestion was stopped with formic acid, and samples were vortexed and centrifuged (800 xg, 20 min) prior to supernatant desalting using spin C18 columns.

#### Mass spectrometry data acquisition

Samples were reconstituted in 0.1% formic acid and were analyzed on a Orbitrap Fusion Lumos with an EASY-Spray nanosource coupled to a nano-UPLC system (EASY-nanoLC 1000 liquid chromatograph) equipped with a 50-cm C18 column (EASY-Spray; 75 μm id, PepMap RSLC C18, 2 mm particles, 45 °C). Chromatographic gradients were started at 5% buffer B with a flow rate of 300 nL/min and gradually increased to 22% buffer B in 79 min and to 32% in 11 minutes. After each analysis, the column was washed for 10 min with 95% buffer B (Buffer A: 0.1% formic acid in water. Buffer B: 0.1% formic acid in acetonitrile). The mass spectrometer was operated in data-dependent acquisition mode, with full MS scans over a mass range of m/z 350-1500 with detection in the Orbitrap (120K resolution) and with auto gain control (AGC) set to 100,000. In each cycle of data-dependent acquisition analysis, following each survey scan, the most intense ions above a threshold ion count of 10,000 were selected for fragmentation with HCD at normalized collision energy of 28%. The number of selected precursor ions for fragmentation was determined by the “Top Speed” acquisition algorithm (max cycle time of 3 seconds), and a dynamic exclusion of 60 s was set. Fragment ion spectra were acquired in the ion trap with an AGC of 10,000 and a maximum injection time of 200 ms. One of the samples, BS04, could not be profiled for technical reasons. All data were acquired with Xcalibur software v4.1.

#### Mass spectrometry protein expression quantification

The MaxQuant software suite (v1.6.2.6) was used for peptide identification and label-free protein quantification^91^. The data were searched against the Uniprot human database with decoy entries (as of April 2018, 42,518 entries). A precursor ion mass tolerance of 4.5 ppm at the MS1 level was used, and up to two missed cleavages for trypsin were allowed. The fragment ion mass tolerance was set to 0.5 Da. Oxidation of methionine, protein acetylation at the N-terminal were defined as variable modification; whereas carbamidomethylation on cysteines was set as a fixed modification. The minimum number of razor and unique peptides (“min. razor + unique”) for a protein group to be considered as identified was set to 1. Identified peptides and proteins were filtered using a 5% FDR. The match between runs algorithm was activated with a tolerance of 0.7 min for the matching time window and 20 min for the alignment time window. The mass spectrometry proteomics data have been deposited to the ProteomeXchange Consortium via the PRIDE^92^ partner repository with accession number PXD011626.

### Pre-processing and differential expression analysis of proteomics data

From the initial set of 649 quantified protein expression profiles on 20 samples, we discarded 26 whose protein identifier could not be mapped to one of the 25,221 genes profiled by RNA-seq. From the remaining 624 proteins, we selected 303 with protein quantification values in at least 6 out of the 20 samples. We further discarded 48 proteins whose protein quantification values were based on one single peptide, leading to a final protein quantification data matrix of 245 proteins by 20 samples. Raw protein quantifications were normalized using the vsn^93,94^ package. A complete normalized matrix of protein expression values was obtained by imputing missing quantifications with a multivariate method^95^ based on the expectation-maximization (EM) algorithm implemented in the function impute.wrapper.MLE from the R package *imputeLCMD*. Using this complete and normalized matrix of protein expression values as an input to the edgeR^80^ and *limma-trend*^85^ pipelines, we produced the MDS plot in Figure 1d and conducted a protein differential expression analysis with linear models shown in Figure 2c. The design matrix included the FIR status of infants as the main explanatory variable and sex as a covariate.

### Functional enrichment analysis with the Gene Ontology database

We performed functional enrichment analysis with the GO database using the package GOstats^96^. More concretely, we used the biological process ontology and the conditional hypergeometric test to account for dependencies derived from the GO hierarchical structure. Under this procedure, the GO hierarchy is traversed towards the root, applying a one-tailed Fisher’s exact test between the genes in the GO term and the DE gene set at hand. When a GO term is considered to be significantly enriched by the DE gene set, its genes are removed from parent GO terms before testing them. To consider a GO term as significantly enriched we used a *P* value cutoff of 0.01, a minimum OR of 1.5, a minimum of five and a maximum of 300 genes in the GO term. The gene universe was defined by the entire set of 25,221 profiled genes by RNA-seq. To report only the most robustly enriched biological processes, the final set of significantly enriched GO terms was selected by first using a multiple testing correction of FDR < 10% on the whole set of tested GO terms, then by further selecting GO terms with OR > 1.5 and a minimum number of five genes enriching the GO term.

### Analysis of T cell receptor excision circles

TREC data were obtained using the EnLite™ Neonatal TREC kit (PerkinElmer, Turku, Finland), which is a combination of PCR-based nucleic acid amplification and time-resolved fluorescence resonance energy transfer (TR-FRET) based detection. EnLite™ simultaneously detects two targets, TREC and beta-actin, the latter used as an internal control for monitoring specimen amplification in each test. The assay involves DNA elution from 1.5mm DBS punches, amplification and hybridization with target-sequence specific probes, and quantifies TREC levels by measuring probe fluorescence with the Victor Enlite™ fluorometer (PerkinElmer). A full calibration curve with blanks and three DBS calibrators was run in triplicate on each plate. A low-TREC control, a no-TREC control, a high-TREC control, and a blank paper disk (containing no blood), were used as quality controls for each plate. Raw fluorescence counts, measured at 615 nm, 665 nm, and 780 nm, were processed by the EnLite™ workstation software to produce corrected fluorescence counts. TREC and beta-actin copies/μL were predicted from corrected fluorescent counts by the EnLite™ workstation software using an unweighted linear regression model on the ArcSinh transformation of the DBS calibrator copies/μL response.

The values for the number of TREC copies/μL are sensitive to technical effects associated with the run in which they are obtained; for this reason, we generated technical replicates for the same individuals in different runs. Because of limited DBS availability, we could not produce more than one TREC read-out for all individuals and not all individuals with multiple readouts had the same number of replicated measurements. For this reason, we used a linear mixed-effects model to test the effect of FIR on the TREC read-out, using the newborn identifier as a random effect variable to account for replicated measurements across individuals. We used the logarithm of the corrected fluorescent counts as TREC read-out instead of the predicted number of copies/μL, as these more accurately reflect the measurements made by the instrument and its technical biases. In this linear mixed-effects model, the FIR stage was the main explanatory variable, and aside from the random effect of the newborn, other covariates that entered as main factors in the model were sex, maximum lymphocyte count during the first postnatal week, and a batch indicator variable identifying the run of the TREC measurement. This linear mixed-effects model, as well as a null one without the FIR stage, were fitted to the data using the package *Ime4*^97^.

A chi-squared test with one degree of freedom between the two models gave a *P* value=0.0432, thus rejecting the hypothesis of no FIR effect at a significance level < 0.05. Adjusted and corrected fluorescent counts on a logarithmic scale (shown in Figure 4a) were obtained after removing the effect of the previous covariates using a linear model.

## Data Availability

The transcriptomics and clinical data reported in this paper are available through the European Genome-phenome Archive (EGA) under accession number EGAS00001003635. The mass spectrometry proteomics data are available through the PRIDE repository under accession number PXD011626.

## Acknowledgments

This work was supported by the Spanish MINECO/FEDER (TIN2015-71079-P). The CRG/UPF Proteomics Unit is part of the Spanish Infrastructure for Omics Technologies (ICTS Omics Tech) and is a member of Proteored, PRB3, which is supported by grant PT17/0019 of the PE I+D+i 2013-2016, funded by ISCIII and ERDF. We acknowledge support from the Spanish Ministry of Economy and Competitiveness, “Centro de Excelencia Severo Ochoa 2013-2017” (SEV-2012-0208), “Unidad de Excelencia María de Maeztu” (CEX2018-000792-M), and “Secretaria d’Universitats i Recerca del Departament d’Economia i Coneixement de la Generalitat de Catalunya” (SGR17-595; SGR17-1020), and the Spanish National Bioinformatics Institute - ISCIII (PT17/0009/0014). The authors thank the ELGAN PAD committee, and Nigel Paneth, Sok Kean Khoo, Pete Haak, Madeleine Lenski and Elizabeth Allred, for granting access to the microarray expression and clinical data from the US ELGAN cohort. We would like to thank Aida Andrés, José Aramburu, Sergi Castellano, Alan Leviton and Cristina López-Rodríguez for their critical remarks on various parts of the manuscript.

## Contributors

DC, TC and RC conceived the study. DC, NB, AS, JMG, ES, FC, CR, AN and JLM collected the data. DC and RC analyzed the data. DC, TC and RC drafted the manuscript. All authors revised and approved the manuscript.

## Ethics approval

The study was approved by the Institutional Review Board from the Hospital Clínic de Barcelona (September 16^th^, 2016; ref. HCB/2016/0713). The parents of the analyzed infants provided written, informed consent.

